# Barriers to Health Behavior Change in People with Type 2 Diabetes: Survey Study

**DOI:** 10.1101/2020.11.22.20236190

**Authors:** Oumoukelthoum Mohamdy

## Abstract

**BACKGROUND:** Diabetes mellitus DM (type 2) is one of the major chronic disease that creates burden at the population level. A prevalence of 422 million patients with diabetes worldwide is detected ^1^. UAE population has an increased prevalence of DM; According to International Diabetes Federation (IDF) 17.3% of the UAE population between the ages of 20 and 79 have type 2 diabetes in 2017 ^2^. UAE has been suffering with the health problems that result from obesity and sedentary lifestyle. The purpose of this study is to understand barriers to health behavior among community patients with diabetes mellitus type 2. METHODS: This is a qualitative survey study of 123 adult individuals with type 2 diabetes who are above 18 years old, who participated in a survey that explore their perception about barriers to health behavior change. The survey was distributed online and in 6 health care centers under ministry of health and prevention in the United Arab Emirates. RESULTS: This study finds that the top barrier for behavior change is difficulty to follow healthy schedule (19.4%), workplace condition and timing (14.3%), and laziness (13%). The least barrier was lack of knowledge about healthier behaviors (5.3%). The best way to motivate behavior change was doctor’s advice during regular visit (26.3%), joining support group (18.4%), and receiving electronic tips and reminders on smartphones (15%) CONCLUSION: The outcomes of this study can aid directly in community programs design and implementation, health education campaigns, and health policies. The findings of this study help in understanding real barriers to initiating and sustaining positive health behavior.

## INTRODUCTION

Changing health behavior is an important step in disease prevention and management. Lifestyle-associated diseases are increasing because they are related to individuals’ behavior and the difficulty to influence them. In type 2 diabetes, American Diabetes Association (ADA) guidelines recommends lifestyle modifications such as diet and exercise, as a foundation for DM type 2 management ^3^. Teaching and educating diabetic patients about the importance of behavior change is crucial. In order to successfully change individuals’ health behavior to a healthier one, healthcare providers need to understand the concept of health behavior change and barriers to change. The concept of behavioral influence on the physical health and the appearance of diseases is not a new concept. It has been discussed and implemented long time ago. The term behavioral medicine had been first defined on 1977 on Yale conference ^4^. The term was formally defined and stressed the important of behavioral-science knowledge application in diagnosis, prevention, treatment, and rehabilitation ^4^.

This paper is concerned with application of behavior-science knowledge in prevention; by understanding barriers to health behavior change among community patients to prevent development of diabetes and use knowledge gained as a guide for designing health promotion programs.

The process of behavior change is controlled by many factors, according to research paper by Robinson et al.^5^ examining facilitators and barriers to the health promotion practice:

“from an interpersonal health behavior perspective, social cognitive and/or learning theory is informative based on its assertion that behavior is a product of the interaction between individual and environmental factors; and therefore, behavior change requires supportive beliefs, training and/or skills, incentives and/or reinforcements, and social and physical environments” ^5^

While the government is spending a lot of resources aiming to reduce the burden of diabetes and other non-communicable diseases (NCDs); diabetes still representing an endemic in the area and the number of cases is still high. In 2015, one million people were reported as having diabetes in the UAE ^6^. The population compliance and engaging on the wide scale programs and services remain a challenge. Healthcare providers’ role especially in primary care needs to expand and cover techniques like behavior change to influence individuals’ decision. On the other hand, there is little research efforts done on this topic to address the problem of poor compliance to health promotion and disease prevention programs. There are studies done in the UAE to explore barriers to certain health behaviors in specific disease population. A research study done by Al-Kaabi, Al-Maskari et al.^7^ on diabetic patients to explore barriers to activities among type 2 diabetic patient found that, cultural issues, especially pertaining to women, represents main barrier to physical activity. Other barriers include the shape of traditional clothes, for both genders, that hide the excess weight and reduce the motivation to physical activity^7^.

Another study that was conducted on seven Arab countries (including UAE) to highlight perceived barriers to healthy eating and physical activity among adolescences ^8^. The study found number of barriers to be prominent among adolescences, barriers to healthy eating were lack of information on healthy eating, lack of motivation to eat healthy diet, and not having time to prepare or eat healthy food^8^. Lack of time was a common barrier for both healthy eating and physical activity among the seven countries.

Barriers to behavior change can be contributed to many factors. It could be – but not limited to-social, psychological, environmental, and financial. A study done on Canada on patients with diabetes to address financial barriers to care found that the main aspects which represent financial barriers were medications, diabetes supplies, and healthy food^9^. Exploring these barriers allowed the patients to seek healthcare providers aid in overcoming these barriers; demanding more thoughtful care and effective strategies to help reducing the financial barriers.

Management of diabetes type 2 revolves around many techniques, currently the most accepted method is self-management. Strategies like lifestyle modifications including diet and physical activity are the main focus of self-management. In order to achieve these goals, it is not enough to educate the patients only but also to monitor their progress and understand what holds them back from meeting their health goals.

Some studies deeply explored barriers to health behavior change and identified what initiates a health behavior and what sustains it. A study by G. Volpp and Mohta founds that Improved access to preventive care is the best strategy for initiating a health behavior whereas in-person social support is the best for sustaining the healthy behavior ^10^. The study involves 775 participants in patient engagement survey done to explore what strategies initiate behavior change and what sustain the change. For long term effects and useful behavior change, the best strategy is in-person social support.

## METHOD, SAMPLE AND DATA COLLECTION

### Method

This is a qualitative survey study. A bilingual online survey (Arabic and English) has been distributed to community patients who have type 2 diabetes [Appendix -1]. Through Ministry of Health and Prevention (MOHAP) media and communication center, the survey was sent to all MOHAP users through email. The survey was also distributed along with the electronic consent [Appendix – 2] in health centers under MOHAP which has NCD clinics, diabetes clinics, and chronic disease community patients. This research has undergone Institutional Review Board IRB process from two committees: MOHAP research ethics committee in UAE and IRB in Arizona State University ASU, USA. MOHAP research ethics committee had approved the study protocol to be conducted (Reference no.: MOHAP/DB-REC/MJJ/No.20/2019).

The study was exemption granted from IRB in Arizona State University (IRB ID: STUDY00010026). The IRB determined that the protocol is considered exempt pursuant to Federal Regulations 45CFR46 (2) Tests, surveys, interviews, or observation on 4/23/2019.

### Sample

The population of this study is community adults’ patients with types 2 diabetes in the UAE. The ideal sample size was calculated considering population size of 1 million (based on IDF report last report in 2017 ^2^). Confidence Level of 90% and margin of error 7.4% were used. The minimum acceptable sample size is 100. Convenient sampling method was followed for collecting data for this research. A total number of 151 responses were recorded, but only 123 samples provided completed questionnaires. The uncompleted surveys were destroyed and not used in the study. 63 responses were collected online, while the other 60 responses were collected from health centers during patients’ regular visits. In the case of 60 responses which were collected in health centers, the process of data collection was handled by the nursing team in the health care center. Any patient with type 2 diabetes who visited their community health center in the period from 26^th^ of April 2019 till 10th of May 2019 was asked to participate in the survey. The sample gender distribution of the 123 participants is: 27 males and 96 females. The age group of the sample varies from 18 years to 74 years with 54.8% of the participants in the age group of 34 to 54 years [Table-1]. The education level of the sample represents large group of educated patients, the majority has a bachelor’s degree as their highest education [Figure-1]. The participants were provided with an electronic consent containing brief description of the study and its importance in planning health programs and strategies for the management of DM type 2.

**Fig. 1.**
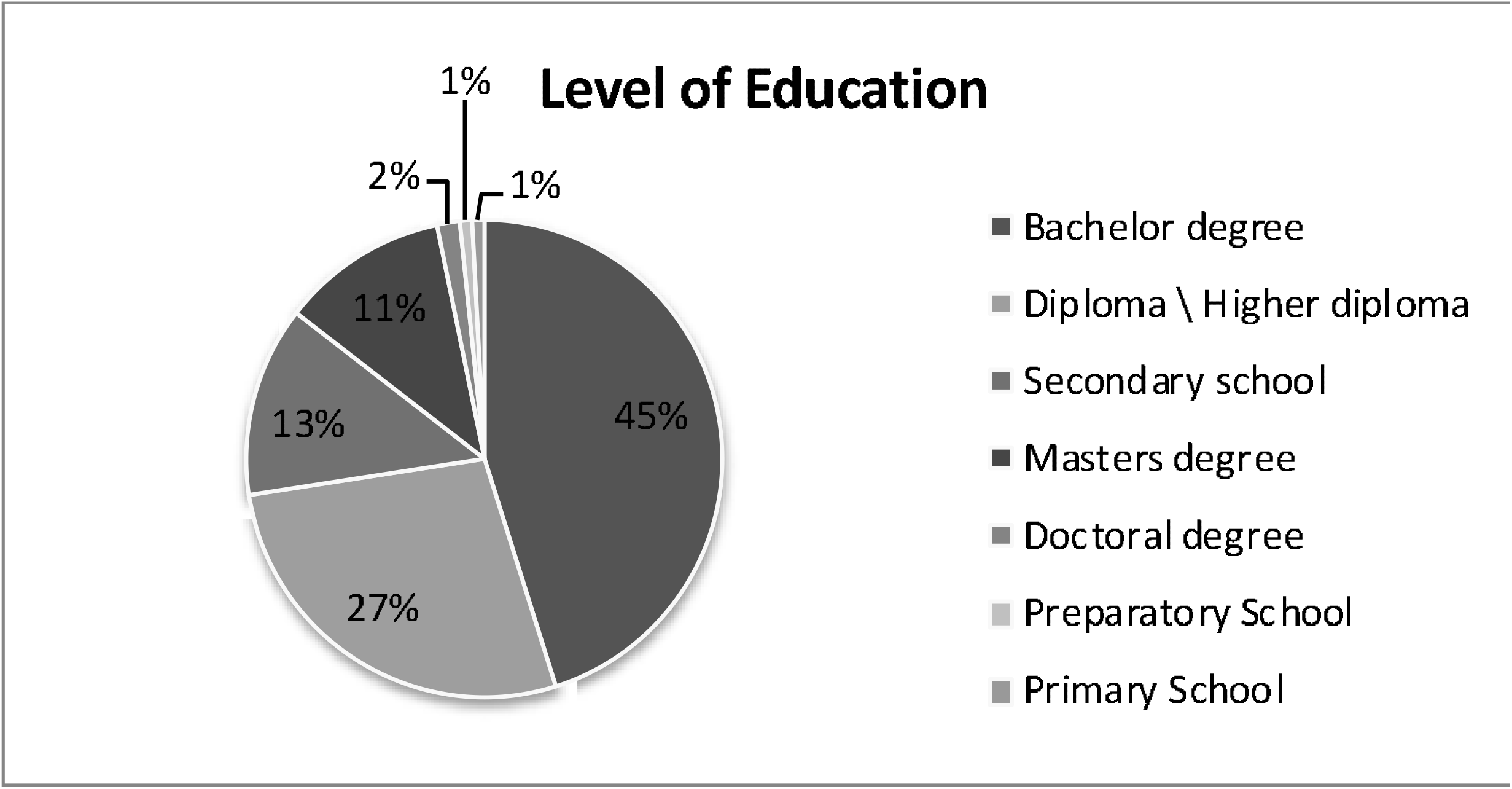
Level of education.

The inclusion criteria are: 1) being diagnosed with type 2 diabetes, 2) above 18 years old, 3) living in the UAE, and 4) not hospitalized during the time of taking the survey. The exclusion criteria are not meeting the inclusion criteria.

### Questionnaire

The survey questions were created to explore the real barriers to health behavior change by obtaining the point of view of patients with DM. In this study, barriers refer to any obstacles of any nature that prevent people with type 2 diabetes from changing their health behavior to a healthier one as instructed by their health providers. The questions contain inquiry about patients’ perception about barriers to behavior change and effective ways to motivate behavior change. 10 questions about participants demographics, history of disease diagnosis, barriers to behavior change, and motivators to behavior change from their personal experience are the main topics in the survey. [Appendix-1]. The last question focuses on actual behavior change experience in which participants succeed in changing their behavior permanently and what was the main motivator to change.

## DATA ANALYSIS

The raw data was collected and recorded by Qualtrics software for online surveys. After completion of data collection, the data was imported to Microsoft Excel to be analyzed. The analysis includes summary statistics for participants demographics and figures and graphs with the percentages for other survey questions.

## RESULTS

The answers to survey questions were analyzed and displayed using bars and graphs. The population characteristics are summarized on Table-1 and Figure-1. The participants’ duration of DM diagnosis is shown on [Figure-2]. The duration of being diagnosed with DM differs between the participants, 28.2% of the participants were newly diagnosed with diabetes (less than a year). A question about attempts to change behavior after being diagnosed with DM showed that: 69.3% of participants started changing their behavior to a healthier one, where 22.7% of the participants denied changing their behavior after being diagnosed and 8% can’t remember.

**Table 1:** Age and gender distribution for the study

|  | Male | Female |
| --- | --- | --- |
| 18 to 34 y | 2 | 35 |
| 34 to 54 y | 15 | 52 |
| 55 to 74 y | 10 | 9 |
| Total | 27 | 96 |
| Percentages | 21.9% | 78.1% |
| Grand Total | 123 |  |

**Fig. 2.**
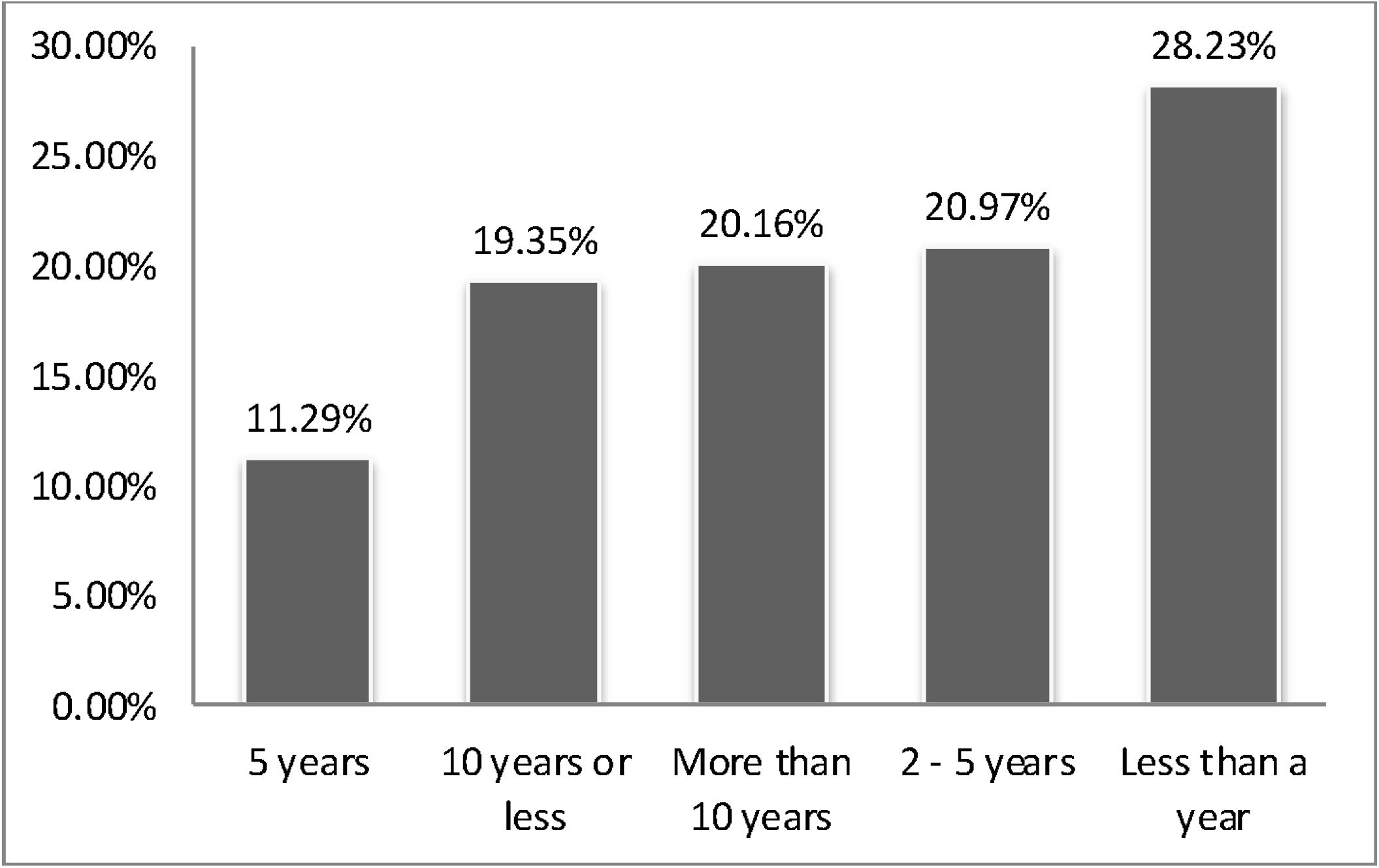
Percentage of participants and their duration of being diagnosed with DM

The core question in the survey is about ratting the top barriers to health behavior change. Participants replied by ratting the top four barriers between nine commonly reported barriers and added other barriers that were not listed in the choices [Figure-3].

**Fig. 3.**
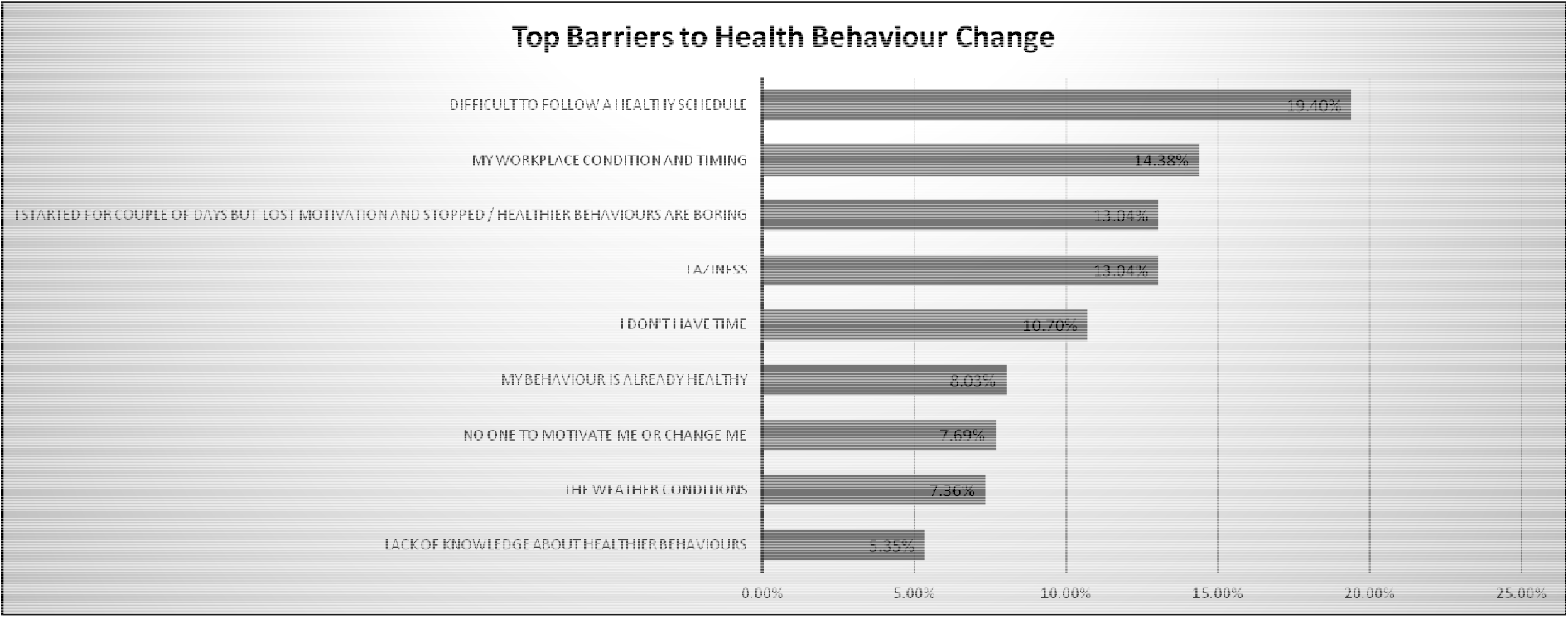
Percentage of top barriers to health behavior change rated by the participants.

The most rated barrier was the answer: “difficult to follow a healthy schedule” 19.4%, on the other hand the least rated barrier was the answer: “Lack of knowledge about healthier behaviors” 5.3%. Other barriers like: workplace condition, time, laziness, and loss of motivation were rated similarly on a range from 14% to 10%. Being already healthy, lack of support, and weather condition were rated less than others on a range from 8% to 7%.

Other barriers that were reported by the participants include: spouse lack of knowledge and temptation of unhealthy food.

After responding to what holds them back from changing their behavior participants reported what are the best ways to motivate behavior change [Figure-4]. The best way was the answer:” doctor’s advice during regular visit”. 26.3% of the participants believed that doctor’s advice to them during their checkups is the best way to motivate them to follow healthy behaviors. The second way is the answer:” joining support group”. 18.4% reported that the support group is essential in motivating behavior change. Motivators like receiving reminders on smartphones, weekly or monthly follow up calls from health facility, and personal health trackers were similar in percentages by 15.4%, 14.3%, and 12.8% respectively. The least way to motivate was the answer:” health session with a community nurse” by 9.4%.

Other motivators that were reported by the participants are: self-education through the internet, body shape, family support, to live longer and fear of diabetes complications.

**Fig. 4.**
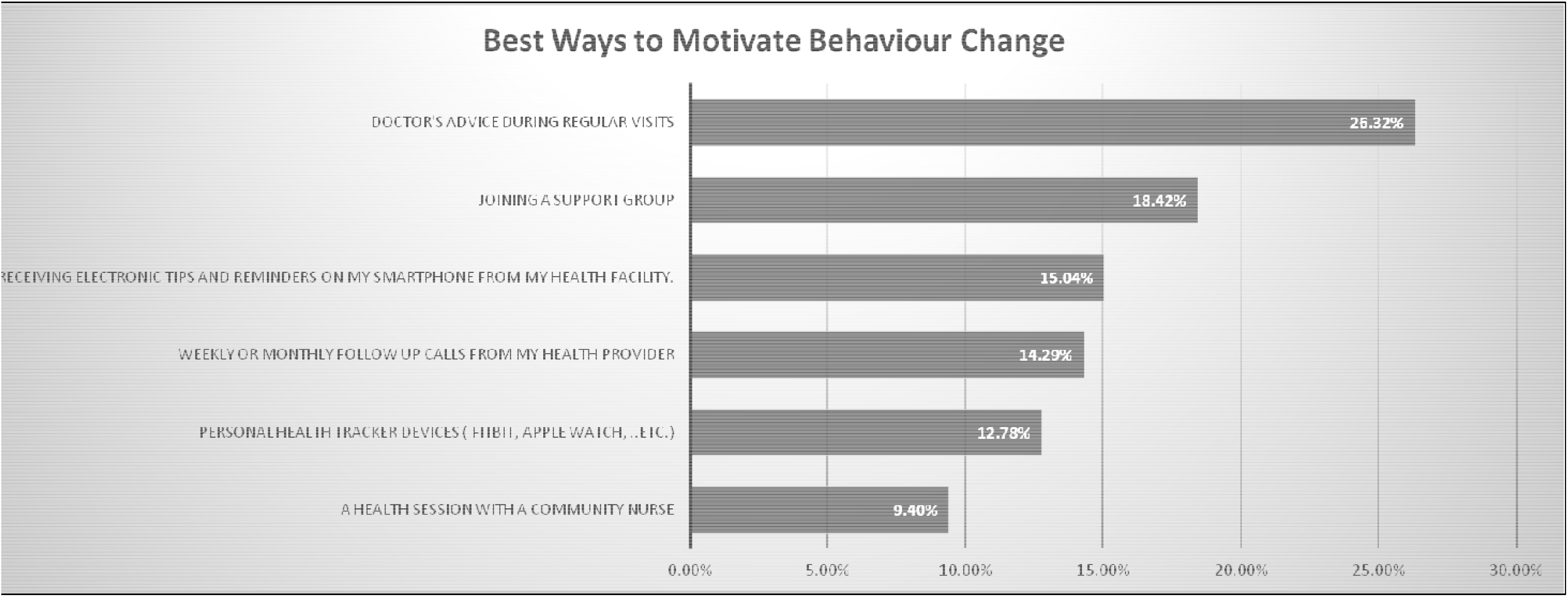
Percentages of effective ways in motivating behavior change.

The last question in the survey was about participants’ experience in permanently changing their behavior. 72.6% of the participants actually succeed in changing their behavior permanently, where 23.4% replied that they never succeed in changing their behavior. Motivators for successfully changing the behavior were: friends support, the presence of nutritionist in the workplace, self-determination, and childbirth.

More than 50% of the participants reported that their family represent the main motivator. The fear of leaving their children and family without support if they didn’t take care of themselves, gave them a reason to change.

## DISCUSSION

This study finds that there are a lot of barriers that stands in the way of patients with diabetes. The sample contains more females than males. The level of education reflects more educated individuals which is attributed to the segment of the sample that received the online survey through MOHAP email. The duration of DM diagnosis differs between the participants, the majority were newly diagnosed and diagnosed within 10 years or less.

This mixture gives the perception of both patients who adapted to the disease management and patients who are newly adapting to DM management. Being diagnosed with DM was the start of behavior change to 69.3% of the participants. The concern of DM complications and comorbidities caused immediate change of behavior.

Barriers to behavior change ratting by the participants reflect important points. The difficulty to follow a healthy schedule was the main barrier in this study. Healthy schedule which includes lifestyle modifications and healthy diets hard to follow. Patients with diabetes receive tips and instructions from their doctor, nurse, and dietician regarding their diet. If following these instructions represents a barrier; health professionals should focus their efforts in strategies to make healthy schedule easier to follow. The active involvement of dieticians and nutrition specialists is important for overriding this barrier. On the other hand, lack of knowledge about healthier behaviors is at the bottom of the list of barriers which draws our attention to important point; patients with diabetes are well informed and have all the needed knowledge about their disease and how to manage it. This finding is consistent with other studies; a study done by Bastin et al.^11^ on barriers to change behavior on obese patients reported that :

> “The vast majority of participants, regardless of the severity of obesity, know they should do and also want to do something to improve their health, but faced a lack of willpower. Thus, the most important thing to consider during an obesity intervention is the lack of motivation to modify health behaviours ….. “

The resources and efforts spent on health education activities should be shifted toward healthy habits implementation and personalized methods to follow healthy schedule. The nutritionists can focus on understanding what each patient needs and tailor the healthy recommendation to their habits.

This study does not test the association between barriers and gender, level of education, and age. However, having almost half the sample (45%) from bachelor’s degree holders indicate that education level may have an effect on reducing the impact of barriers like lack of knowledge (the least reported barrier).

The second top barrier is the workplace condition and timing. Workplace conditions refer to how convenient is the place a patient with diabetes working in to facilitate maintaining health behavior. Long hours, short breaks, and lack of healthy food on the workplace are representing huge challenge for patients with diabetes. To override this barrier, employers should include the healthy habits principles in their agendas like scheduling exercise time during the working hours, offering healthy snacks, and provide nutrition counselor if applicable.

Another way is the presence of nutritionist in the workplace to follow up and measure the progress of employees’ health habits, which was reported to be one of the motivators to behavior change.

Other barriers like laziness, lack of time and loss of motivation are personal factors that can be controlled by the patient and his healthcare provider. These points should be assessed in each patient in order to be addressed if they are representing a barrier. Some participants reported that spouse lack of knowledge is one of the barriers to behavior change. Plans can be done to provide training and health education to family members.

Utilization of principles of behavior change theories and putting it into practice can be very effective in overriding barriers. The theory of planned behavior was used by nurse practitioners to develop a framework that tailored strategies for diabetes self-management^12^. They used the central concept of the theory which is “intention” as the best predictor for behavioral change and start by assessing it very early in the treatment^12^. The paper highlighted the need for high-quality care, which include strategies to address psychological barriers to adhering to diabetes self-management programs^12^.

When analyzing the best ways to motivate behavior change, doctors’ advice during regular visits was the best motivator. Patients tend to value the doctor’s advice and pay more attention to it. This finding is consistent with findings of other studies which linked patient’s adherence to medical care and their trust in the medical profession ^13^. Doctors must maintain good practice in assessing and addressing barriers to health behavior change and develop plans with the help of patients. The second-best way is joining a support group. The role of support group in motivating behavior change and override barriers is tremendous. Support group concept needs to be well established and utilized here in the UAE. Volunteers association and health facilities can establish support groups and recommend it to promote behavior change.

The other motivators can be categorized under the use of technology in changing the behavior: Sending reminders and tips to patients’ smartphones by the health facility, weekly and monthly follow up calls by the healthcare provider, and personal health trackers. These methods were reported by participates to be the effective in motivating behavior change. Weekly or monthly follow up calls are proved to be effective in sustaining behavior change by other studies^11^. This method represents in-person social support.

The least method for motivating behavior change was health session with community nurse. Although the role of nurses in health education and promotion is known to be effective by many studies; this study finds that participants favor other ways of motivation than health session with a community nurse. It is recommended to fully utilize the nurse knowledge and expertise in counseling and dealing with patients who have chronic diseases including diabetes mellitus.

According to WHO recommendation; the fact that nurses are the largest group of health-care providers, they can make an important contribution to efforts to prevent, reduce and treat noncommunicable diseases (NCDs)^14^

Strategies like proper training and introduction of nurse led NCD clinics can have huge impact in changing population health behavior.

## CONCLUSION

The outcomes of this study can aid directly in community programs design and implementation, health education campaigns, and health policies. The findings of this study help in understanding real barriers to initiating and sustaining positive health behavior. Healthcare professionals should work with patients with diabetes by assessing and addressing the top barriers which is facing this type of patients.

### LIMITATION

The study has some limitations in terms of recruiting participants and data collection. Online surveys have the advantage of reaching out more responses but result on bias; only individuals who has access to the internet and are literate enough to answer the survey had the chance to participate. For this type of research, the best method of data collection is face-to-face interactive interview to capture more authentic responses and avoid reply bias.

For future research, it is recommended to conduct more studies on the area of behavioral sciences and its implication in the UAE. Designing interventions for health promotion program using the findings of this study is recommended. Strategies to Involve all health professionals especially dieticians in changing the behavior of patients with diabetes. More Nursing research to study utilization of nurse-led intervention in changing behavior is recommended.

. It is also important to use the technology and innovative methods to motivate, initiate and sustain healthy behavior.

## Data Availability

All Mentioned Data is available

## ACKNOWLEDGMENT

This research was supervised by Professor Kristen Will : (MHPE, PA-C Director, Executive and Continuing Education, Clinical Assistant Professor, Science of Health Care Delivery Arizona State University | College of Health Solutions) as a principal investigator. This is to extend special thanks and appreciation for her guidance and expertise which added to the quality of this research study.

This is also to extend many Thanks and appreciation to the nursing team in Sharjah primary health care centers, ministry of health and prevention for supporting and participating in the process of data collection.

## Statement of Ethics

This research has undergone Institutional Review Board IRB process from two committees: MOHAP research ethics committee in UAE and IRB in Arizona State University ASU, USA. MOHAP research ethics committee had approved the study protocol to be conducted (Reference no.: MOHAP/DB-REC/MJJ/No.20/2019).

The participants were provided with an electronic consent containing brief description of the study and its importance in planning health programs and strategies for the management of DM type 2 prior to answering the survey.

## EXEMPTION GRANTED

Kristen Will

CHS: Executive Education

602/496-0808

Dear Kristen Will:

On 4/23/2019 the ASU IRB reviewed the following protocol:

|  |  |
| --- | --- |
| Type of Review: | Initial Study |
| Title: | Understanding Barriers to Health Behaviour Change: Perception of Patients with Type 2 Diabetes in United Arab Emirates UAE |
| Investigator: | Kristen Will |
| IRB ID: | STUDY00010026 |
| Funding: | None |
| Grant Title: | None |
| Grant ID: | None |
| Documents Reviewed: | • Survey Questions AR.pdf, Category: Recruitment Materials;<br>• citiCompletionReport7729978.pdf, Category: Other (to reflect anything not captured above);<br>• translation form.pdf, Category: Translations;<br>• Study Protocol(Revised), Category: IRB Protocol;<br>• Dual Role.pdf, Category: Other (to reflect anything not captured above);<br>• Consent form(Revised).pdf, Category: Consent Form;<br>• Survey Questions EN.pdf, Category: Recruitment Materials;<br>• Wizard.pdf, Category: Other (to reflect anything not captured above); |

The IRB determined that the protocol is considered exempt pursuant to Federal Regulations 45CFR46 (2) Tests, surveys, interviews, or observation on 4/23/2019.

In conducting this protocol you are required to follow the requirements listed in the INVESTIGATOR MANUAL (HRP-103).

Sincerely,

IRB Administrator

cc: Oumoukelthoum Mohamdy Oumoukelthoum Mohamdy

### Ministry of Health and Prevention

#### Research Ethics Committee

Study Title: Understanding Barriers to Health Behaviour Change: Perception with type 2 Diabetes in the UAE

Subject : Approval Reference No: MOHAP/DXB-REC/MJJ/No.20/2019. Dear Dr. Kristen Will,

Dear Mrs. Oumoukelthoum Mohamdy,

In regards to the above mentioned Study protocol, this is to confirm that on the meeting dated (23 /6/2019), the Ministry of Health and Prevention Research Ethics Committee has reviewed the study protocol as well as all the documents submitted in the submission file from the ethical point of view and has approved the conduct of above mentioned study.

Opinion: Approval

Please find below a list of approved documents:

| Document | Version/date |
| --- | --- |
| Application Form | Ministry of Health and Prevention Application Form-amended version |
| Protocol | Research Summary_- Amended version |
| Information sheet and Informed Consent Form | Electronic Consent - English and Arabic_ Amended version |
| Data Collection | Survey Questions-English and Arabic version _Amended |
| Investigator/s CV | CV/s of Principal Investigator/s |
| GCP Certificate/s | GCP Certificate/s of Principle investigator/s |

The MOHAP Research Ethics Committee is organized and operated according to guidelines of the International Conference on Harmonization and constituted according to ICH-GCP requirements.

**This Ethical approval applies for the following study sites only:**

Wasit Health Center,Oarain Health Center, Batayih Health Center, AIDhaid Health Center, Sharjah Family Health Promotion Center, Hamria Health Center,.A.I Oassimi·Hospital Diabetes ciinic \outpatients.

This approval is subject to the following conditions:

1. The MOHAP research ethics committee approval does not imply that the researcher is granted access to data, medical records or biological samples from the MOHAP health care facilities neither the Private MOHAP licenced health care facilities. Researchers must seek permission and follow the policy and procedure from the concerned directories after the approval from the Research Ethics Committee
2. Please note that it is the Principal Investigator’ s responsibilities, to immediately inform the Committee of any changes in the research protocol and/or the research Methodologies, should the need for those changes arise prior to or during the conduct of this research study
3. The approval is valid for· up to 1year from the date of approval. If the study extends beyond this date, a progress report must be sent to the research ethics committee to renew the approval 30 days prior the expiry.
4. The research ethic committee must be informed when the research has been completed and a copy of the final research report must be submitted for our records.

Yours sincerely

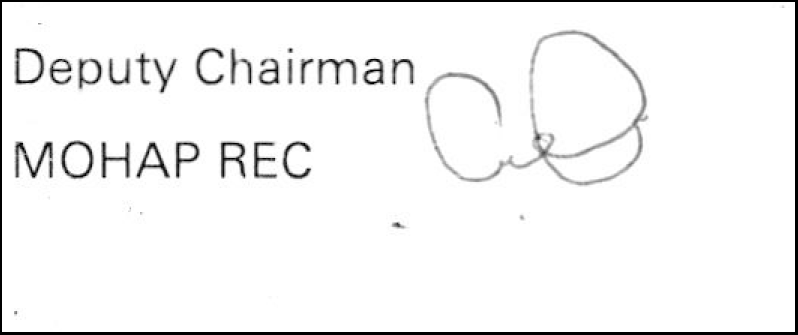

## Consolidated criteria for reporting qualitative studies (COREQ): 32-item checklist

### Domain 1: Research team and reflexivity

Personal Characteristics

1. Interviewer/facilitator: Which author/s conducted the interview or focus group? Self-reported survey
2. Credentials What were the researcher’s credentials? E.g. PhD, MD Author: Nursing Unit Manager with MS in health care delivery Principal Investigator: PhD
3. Occupation What was their occupation at the time of the study? Author: Nursing Unit Manager with MS in health care delivery, Ministry of Health and Prevention, United Arab Emirates, Primary Health care Sector. Principal Investigator: Director, Executive and Continuing Education Clinical Assistant Professor, Science of Health Care Delivery Arizona State University | College of Health Solutions
4. Gender Was the researcher male or female? Females
5. Experience and training What experience or training did the researcher have? CITI program Training

### Relationship with participants

6. Relationship established Was a relationship established prior to study commencement? No
7. Participant knowledge of the interviewer No What did the participants know about the researcher? Purpose of the research, academic background, name and gender, occupation. research
8. Interviewer characteristics What characteristics were reported about the interviewer/facilitator? Intrserted in this topic as it is a main field in my work (Community nursing)

### Domain 2: study design

Theoretical framework

9. Methodological orientation and Theory What methodological orientation was stated to underpin the study? content analysis, observations from practice. Participant selection
10. Sampling How were participants selected? Specific disease group : adults with type 2 diabetes.
11. Method of approach How were participants approached? face-to-face, email
12. Sample size How many participants were in the study? 151 (total) 124 (completed surveys)
13. Non-participation How many people refused to participate or dropped out? Reasons? 27, uncompleted surveys.

### Setting

14. Setting of data collection Where was the data collected? clinic, online
15. Presence of non-participants Was anyone else present besides the participants and researchers? Family members, healthcare providers at the assessment room
16. Description of sample What are the important characteristics of the sample? Mixture of males and females, age range from 18 till 55 years

### Data collection

17. Interview guide Were questions, prompts, guides provided by the authors? Was it pilot tested? provided by the author, first time used.
18. Repeat interviews Were repeat interviews carried out? If yes, how many? no
19. Audio/visual recording Did the research use audio or visual recording to collect the data? No
20. Field notes Were field notes made during and/or after the interview or focus group? no
21. Duration What was the duration of the interviews or focus group? 4-6 mintues
22. Data saturation Was data saturation discussed? no
23. Transcripts returned Were transcripts returned to participants for comment and/or correction? no

### Domain 3: analysis and findingsz Data analysis

24. Number of data coders How many data coders coded the data?N/A
25. Description of the coding tree Did authors provide a description of the coding tree? N/A
26. Derivation of themes Were themes identified in advance or derived from the data? N/A
27. Software What software, if applicable, was used to manage the data? Microsoft Excel
28. Participant checking Did participants provide feedback on the findings? no

### Reporting

29. Quotations presented Were participant quotations presented to illustrate the themes / findings? Was each quotation identified? e.g. participant number No
30. Data and findings consistent Was there consistency between the data presented and the findings? yes
31. Clarity of major themes Were major themes clearly presented in the findings? yes
32. Clarity of minor themes Is there a description of diverse cases or discussion of minor themes? yes

